# # AstraZeneca vaccine disinformation on Twitter

**DOI:** 10.1101/2021.04.08.21255107

**Authors:** Dariusz Jemielniak, Yaroslav Krempovych

## Abstract

We analyzed 50,080 tweets about # AstraZeneca in English from 2021. We found that the news most common in the frequently retweeted tweets abound in negative information, and in many cases come from media sources well-known for disinformation. Also, we found that RT, a Russian state-sponsored news website, as well as Al Arabiya, a Saudi-owned news website, are frequently retweeted with information about the vaccine. Our analysis identified large coordination networks involved in political astroturfing and vaccine diplomacy in South Asia but also vaccine advocacy networks associated with European Commission employees. Our results show that Twitter discourse about # AstraZeneca is filled with disinformation and bad press, and may be distributed not only organically by anti-vaxxer activists, but also systematically by professional sources.

**Highlights:**

- Most commonly retweeted media news links abound in negative information
- Well established disinformation media sources are retweeted more
- State coordinated networks are active in # AstraZeneca astroturfing

## Introduction

Anti-vaxxer discourse in social media is particularly prone to disinformation, additionally more dangerous in the times of the COVID-19 pandemic [1,2]. It is also often amplified by social bots, which disproportionately more often retweet content [3].

As the rollout of different vaccines continues, the problem exacerbates. A particularly interesting case is the AstraZeneca vaccine, due to its media coverage related to the allegedly lower comparative efficacy, tensions over EU-UK exports, as well as an alleged link to very rare cases of blood clots [4].

In order to study the social media discourse on AstraZeneca, we decided to focus on media coverage of # AstraZeneca, and the most retweeted links and tweets. In the second stage, we have also analysed co-tweet networks and bot activity related to the discourse.

## Method

We conducted a Thick Big Data [5] analysis of the AstraZeneca hashtag on Twitter. We collected 221,922 tweets with # AstraZeneca from January 1, 2021, to March 22, 2021. We focused on 50,080 of these tweets, which were in English. Furthermore, we established the final URLs linked in these tweets, by extracting them from popular link shorteners (such as bit.ly, ow.ly, etc.).

We focused on two periods: before and after March 7, 2021 -the date when Austrian authorities took a precautionary step to suspend vaccinating with a batch of AstraZeneca’s COVID-19 vaccine, after the death of one person and reported illness of another after the shots.

Furthermore, we studied which media were the most often linked to among the tweets that were retweeted more than 10 times for both periods. We were interested in discovering whether there was a change from mainstream to alternative sources after the allegation of clots was publicized. We also analyzed the most liked and most retweeted tweets qualitatively.

In the second stage of our investigation, we concluded Coordination Detection analysis [6] to identify cooperative efforts to propagate some tweets. To detect coordination, we conducted similarity analysis of tweets corpuses using the Min-Hash [7] local-sensitive hashing method, which has allowed us to identify similar items based on the Jaccard similarity between sets of strings’ hashtag n-grams. For this analysis, we have adopted a 0.8 Jaccard similarity threshold.

To project each tweet’s coordination network, we drew an edge between two accounts with matching tweet text corpus using co-occurrence analysis functionality from the quanteda package for R [8]. In the output, we have reached a graph with forty-two hundred nodes representing tweets with a similar or identical corpus which accounted for more than eleven thousand unique Twitter user handles.

We have also performed bot identification using a Botometer machine learning platform that computes a bot likeliness ranking with low scores indicating likely human accounts and high scores indicate possible bot accounts. Botometer measures the score by comparing an account against tens of thousands of labelled inputs in its database [9], considering prior coordination detection analysis that we performed and upon review of the body of works utilizing this tool in a similar context [10], we set a Complete Automation Probability score to 0.76 and above.

## Results

Before March 7, in the top 10 linked sources among the tweets retweeted more than 10 times 4 were established Western news media, such as AFP (# 18,806 most popular website according to Alexa traffic ranking), Politico.eu (# 20,905 in Alexa), Telegraph (# 1,730 in Alexa), or the Guardian (# 172 in Alexa) -see Figure 1.

**Figure 1:**
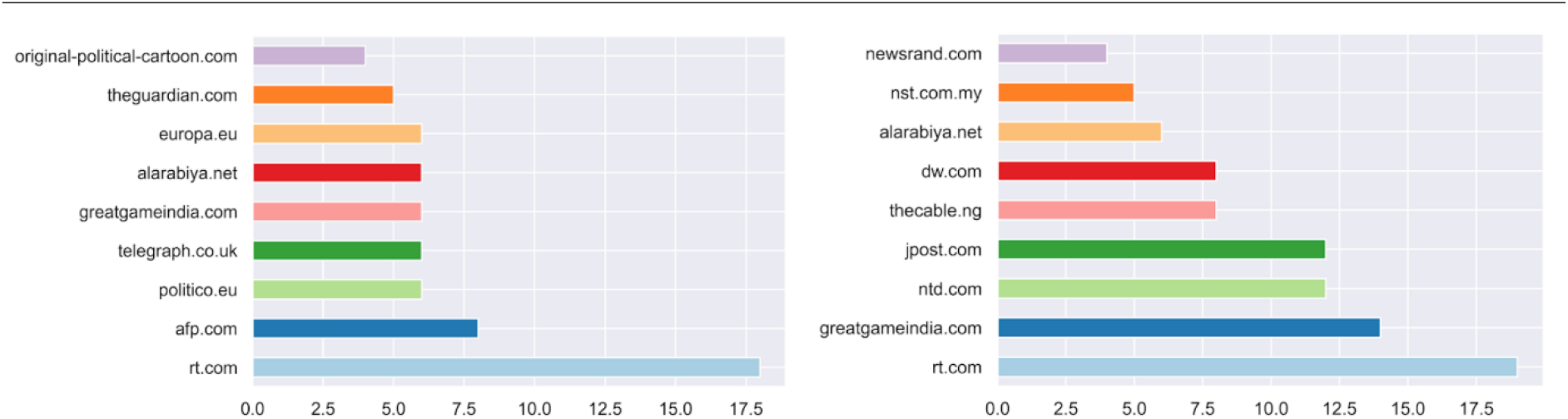
Media linked to in tweets retweeted more than 10 times before (left) and after (right) the news of alleged blood clots

After March, 7 NewsRand, a relatively unpopular Nigerian news website (# 126,271 in Alexa) easily won with the aforementioned mainstream outlets, which all dropped out from the top ten. Other high positions were taken by less surprising, well-established popular media such as the German DW (# 650 in Alexa), the Jerusalem Report (# 5,550 in Alexa), The Cable from Nigeria (# 11,526), and Malaysian New Straits Times (# 20,625 in Alexa), or NTD (a Chinese television from New York, # 28,189 in Alexa).

Other discoveries are more interesting. First, GreatGameIndia (# 126,482 in Alexa), an Indian website, which is known for disinformation, conspiracy theories, and in particular COVID-19 fake news [11], was number 5 before March, and number 2 in the period after.

Second, the position of Al Arabiya (# 2,108 in Alexa), a Saudi-sponsored, propagandist Arabic news channel [12], was relatively stable in both studied periods (6th and 7th respectively).

Third, RT (# 312 in Alexa), formerly known as Russia Today, had a clear lead in both periods. Russia Today is a state-owned media outlet well known for disinformation [13,14] and supporting Russian diplomatic goals as an information warfare tool [15].

Even though links to RT are not contained in the top 100 most retweeted tweets (requiring 73 or more retweets), they are retweeted frequently enough in smaller numbers to nevertheless bring RT to the leading position among tweets with over 10 retweets overall. We believe that this may suggest a non-organic propagation of news by an orchestrated group of professionals.

When tweets retweeted more than 2 times are considered, Al Arabiya takes the lead in both periods, and RT is third in the earlier period and second in the latter one. For tweets retweeted more than 3 times Al Arabiya and RT are number one and two respectively in both periods, and their tweets constitute 31% of tweets from the top 10 most popular sources in the first period and 35% in the second. A qualitative analysis of RT links shows that a large part of them are negative in describing the AstraZeneca vaccine.

Outside media links, the most retweeted (2656 retweets as of March, 26) tweet overall from the first period was one by Robert Kennedy Jr, a known anti-vaxxer advocate, whose account on Instagram was terminated in February 2020 because of COVID-19 disinformation [16]. The tweet was discrediting AstraZeneca vaccine as “controversial”, “heavily invested in by Bill Gates”, and “being rejected over widespread concerns”. The most retweeted (2015 retweets as of March, 26) tweet from the second period was one by Disclose.tv, a well-established disinformation site [17].

Tweet coordination analysis has revealed 10,728 instances in the coordination carried out by 1137 unique handles, out of which 2,278 and 616 unique handles are related o automatic bot accounts, according to Botometer Complete Automation Probability score of 0.76 and above. The largest coordination network had 451 co-ordination instances by 111 accounts, out of which 74 scored above our threshold on the Botometer. The second-largest network consisted of 37 accounts responsible for 47 coordination instances; out of these 37 accounts 13 accounts can be considered automated -see Figure 2.

**Figure 2:**
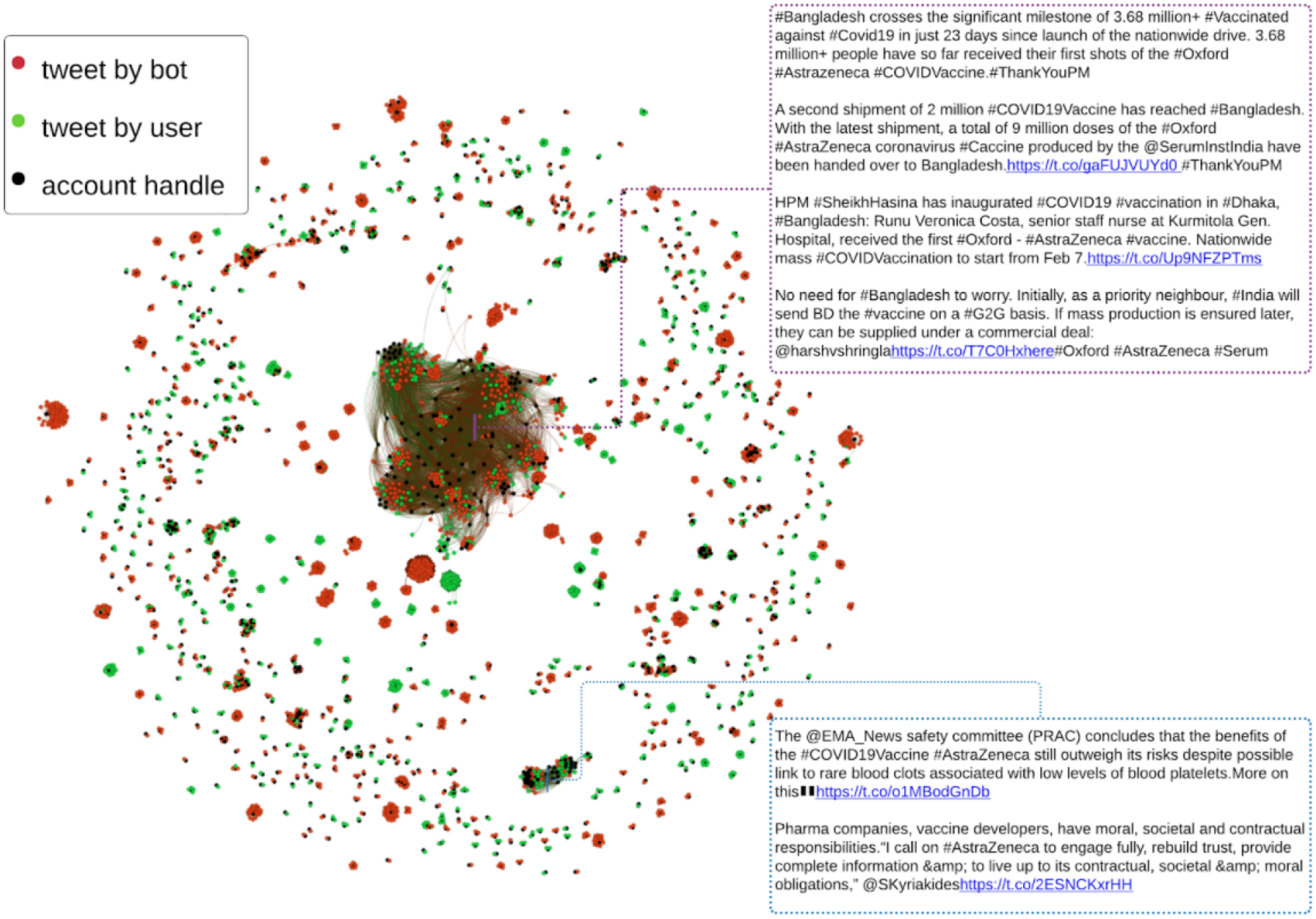
Co-tweets networks discovered in the dataset

While analyzing tweet contents of the first coordination network, we discovered that it connects accounts that tweet about current vaccination situations in Bangladesh. The coordination network contained tweets praising Bangladesh Prime Minister Sheikh Hasina’s actions in battling the coronavirus pandemic and used the hashtag # ThankYouPM. In some contexts, the same hashtag is often used on Twitter to refer to India’s prime minister -Narendra Modi.

Another topic presented in this coordination network is the relation between Bangladeshi and Indian states in large vaccine donations by the latter to Bangladesh. It is essential to mention that Bangladesh’s neighboring country -India is one of the world’s largest vaccine producers. Indian government actively explores opportunities associated with control over vaccine distribution to strengthen its neighborhood ties through vaccine diplomacy [18]. According to the analysis prepared by Oxford Analytica -Bangladesh and Myanmar are the biggest recipients of vaccine donations from India [18].

In both cases presented above, automated bot account are engaged in an activity of political astroturfing: “[…] a centrally coordinated disinformation campaign in which participants pretend to be ordinary citizens acting independently, has the potential to influence electoral outcomes and other forms of political behavior“ [19]. In our coordination analysis, astroturfing activities involved a co-tweet coordination network. Automated bot accounts spread identical or very similar texts to amplify political message reach, in this case -vaccine diplomacy.

When it comes to the second largest coordinated network, the ratio of accounts involved to the number of coordination instances is much smaller while representing only two coordinated messages. Those messages were concerned with official stand of European Medicines Agency’s (EMA) the Pharmacovigilance Risk Assessment Committee (PRAC) regarding safety and efficacy of AstraZeneca vaccine which went under much criticism because of reports suggesting increased risk of blood clots after administering the vaccine in patients [4].

After analyzing the user description and content of their communications, we concluded that this is a network composed predominantly of European Commission’s employee’s accounts and high-level officials, even though 35% of these accounts are considered in this analysis to be automated. Moreover, research of links shared indicated that all the messages connected to the article “Remarks by Commissioner Stella Kyriakides on vaccines” [20]. All these messages used shortened URL from the domain “smh.re” that indicates the use of employee advocacy software Smarp [21]. Considering the facts mentioned earlier, one can safely assume that this network presented a centralized health advocacy communication campaign co-ordinated by the European Commission’s employees to address increased criticism of AstraZeneca vaccines.

### Limitations

For this coordination analysis, we do not take into account networks that share only one handle. However, it is worth mentioning that these networks could be created due to handle sharing, changing an account’s “screen_name”. Given the availability of historical snapshot data about accounts in question, analysis of handle sharing behavior could bring additional perspectives to this research.

## Discussion and conclusions

Our results focus on a short period, and only on one vaccine-related hashtag. Nevertheless, the picture that emerges is deeply troubling. Twitter discourse about # AstraZeneca abounds in disinformation, and reputable media news sources representation is, at best, on par with the disinformation ones, and at worst significantly smaller.

Popular disinformation tweets are spread not only by individual powerful activists and conspiracy websites but also by state-owned media, supported by networks of disinformation farms. Given the fact that Russia has a heavy interest in promoting its own Sputnik-V vaccine, both for economic and political reasons, the activity of RT in posting often negative information about # AstraZeneca may be perceived as part of a larger campaign, potentially aimed at discrediting the vaccine.

At the same time, states like India and Bangladesh foresaw an opportunity in vaccine diplomacy. Results show coordinated astroturfing campaigns with the use of co-tweeting bot networks. In the same time, from another side of the barricades, European Commission’s employees coordinate health advocacy communication promoting the safety of the AstraZeneca vaccines through their accounts.

## Data Availability

data available upon reasonable request

